# Protocol for Systematic Review of Interventions to Improve Sleep Health in Adolescents Living in Low-and-Middle-Income Countries

**DOI:** 10.1101/2023.09.26.23296177

**Authors:** Oluwatosin Eunice Olorunmoteni, Jeremiah Oluwatomi Itodo Daniel, Temitayo Ayantayo, Virginia Guvoeke, Lordstrong Akano

## Abstract

**Introduction:** Adolescence is a transitory phase marked by significant changes. While adolescents are not exempted from the increasing prevalence of sleep-related issues, an urgent need for understanding the factors responsible for sleep problems in this age group and evaluating the possible interventions to mitigate this scourge has risen globally. However, despite facing substantial sleep-related issues, there has been no focus on adolescents in low-middle-income countries (LMICs). This study seeks to serve as a much-needed tool to evaluate and compare the different interventions for adolescents with sleep-related problems in LMICs and provide valuable insights to stakeholders involved with adolescents to improve overall adolescent health.

**Methods and Analysis:** The study will follow the Preferred Reporting Items for Systematic Reviews and Meta-Analysis [PRISMA] guideline 2020. Eligible studies for this review will be identified following an exhaustive literature search on the following databases: PubMed, Embase, Scopus, PsychInfo, and AJOL. Our outcomes will include quantitative measures of sleep health and other health indicators.

Data will be presented qualitatively using tables and diagrams, with narrative synthesis. Two independent reviewers will screen, review, and extract data from studies, with a third reviewer resolving discrepancies. Additionally, we will assess study details, the risk of bias, and the strength of evidence using the Grading of Recommendations Assessment, Development, and Evaluation framework.

**Ethics and dissemination:** This review will be conducted using existing studies; hence, ethical approval will not be required. We aim to present our findings at conferences and publish our manuscript in a peer-reviewed journal for widespread dissemination.

## Introduction

Adolescence encompasses ages 10 to 19, a transitional phase from childhood to adulthood characterized by profound physical, cognitive, and psychosocial changes in which sleep health plays a vital role. (1) It encompasses a multifaceted sleep-wake cycle that adapts to an individual’s social and environmental demands. Sleep patterns hold paramount significance as adolescence progresses, profoundly influencing adolescents’ overall health and well-being. During this transitional phase, adequate sleep and optimal cognitive function are essential for academic success, emotional regulation, and maturation.

The spotlight on adolescent sleep health has intensified globally due to the escalating prevalence of sleep-related issues. (2) These issues include inadequate sleep duration, poor sleep quality, and irregular sleep patterns. The factors contributing to these issues are multidimensional, spanning socio-cultural and economic influences.

Notably, adolescents in LMICs face a disproportionately large burden of sleep problems compared to their counterparts in high-income countries (HICs). They face academic pressures, social obligations, the pervasive influence of electronic devices, inadequate sleep health education, and limited access to healthcare. (3) These can lead to cognitive deficits and other unpalatable outcomes, impeding their career prospects. Various interventions have been executed in response to these issues, encompassing a spectrum of strategies ranging from educational initiatives and lifestyle adjustments to technology-driven approaches. (4–7)

Nevertheless, despite the growing body of research on interventions (8) targeting sleep health in adolescents, conducting a systematic review and meta-analysis remains imperative to comprehensively assess the existing evidence, synthesize findings, and provide valuable insights into the impact of interventions on sleep health in adolescents living in LMICs. This review aims to inform future interventions, policies, and research efforts to enhance adolescents’ well-being and sleep health in LMICs by identifying successful strategies and highlighting areas requiring further attention.

### Study Objectives

- To assess the prevalence of sleep-related problems among adolescents in LMICs
- To review the pattern of sleep disorders among adolescents in LMICs
- To identify factors associated with sleep-related problems in adolescents in LMICs
- To identify and quantify the outcome of interventions successfully addressing adolescents’ sleep health and cognitive function issues in LMICs.

### Research Questions

1. What is the prevalence and pattern of sleep-related problems among adolescents in low- and middle-income countries (LMICs)?
2. What are the various intervention strategies, including educational initiatives, lifestyle adjustments, and technology-driven approaches, that have been implemented to address sleep health issues among adolescents globally, with a specific focus on LMICs?
3. How effective are the interventions in improving sleep health, and the ease of adoption of these strategies
4. How do these interventions improve their sleep health and the link to their quality-of-life outcomes (e.g., cognitive function)

### PICO Framework

#### Population

Adolescents aged 10 to 19 years living in LMICs.

#### Intervention

any intervention on sleep health

#### Comparators

primary research studies on interventions for child and adolescent sleep health vs. studies without interventions

#### Outcomes

Quantitative measure of adolescents’ sleep

## Methods

### Information Sources and Search Strategy

The researchers will identify eligible studies published from inception to date using an exhaustive search strategy across databases like PubMed, Google Scholar, Embase, Scopus, PsychInfo, and AJOL. This systematic review will follow the Preferred Reporting Items for Systematic Review and Meta-Analysis Protocols (PRISMA-P) guidelines.

The researchers will search the titles/abstracts/keywords using these search terms and boolean characters: (Adolescents OR Teenagers OR Youth OR Young adults) AND (LMICs OR Low- and-middle-income countries OR Developing countries OR Economically disadvantaged regions) AND (Sleep disorders OR Sleep disturbances OR Sleep problems OR Insomnia OR Sleep deprivation OR Sleep quality OR Sleep patterns) AND (Factors OR Determinants OR Educational initiatives OR Lifestyle adjustments OR Technology-driven interventions OR Behavioral therapy OR Counseling OR Pharmacological interventions OR Sleep duration OR Sleep quality OR Sleep patterns OR Cognitive function OR Quality of life)

#### PubMed

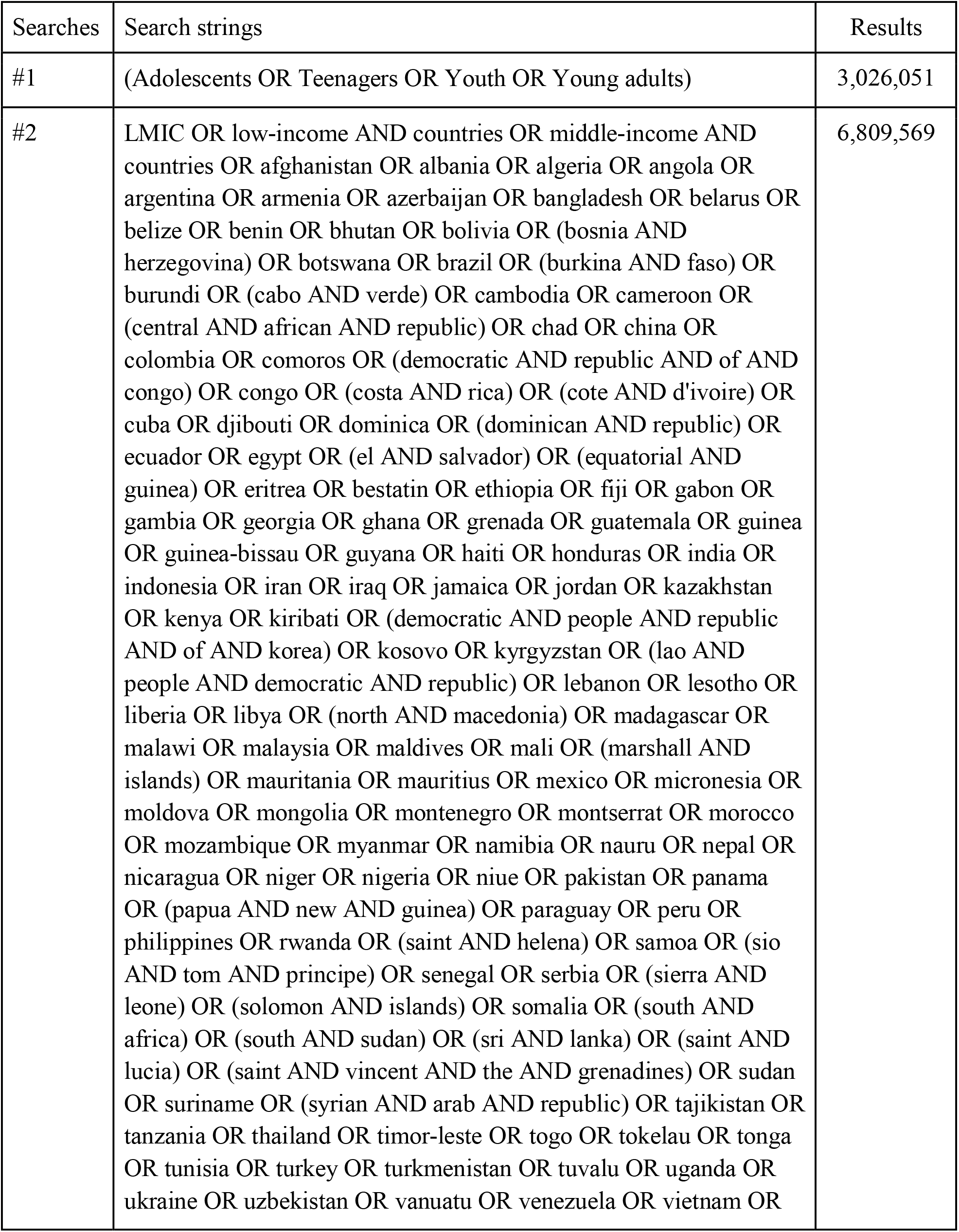
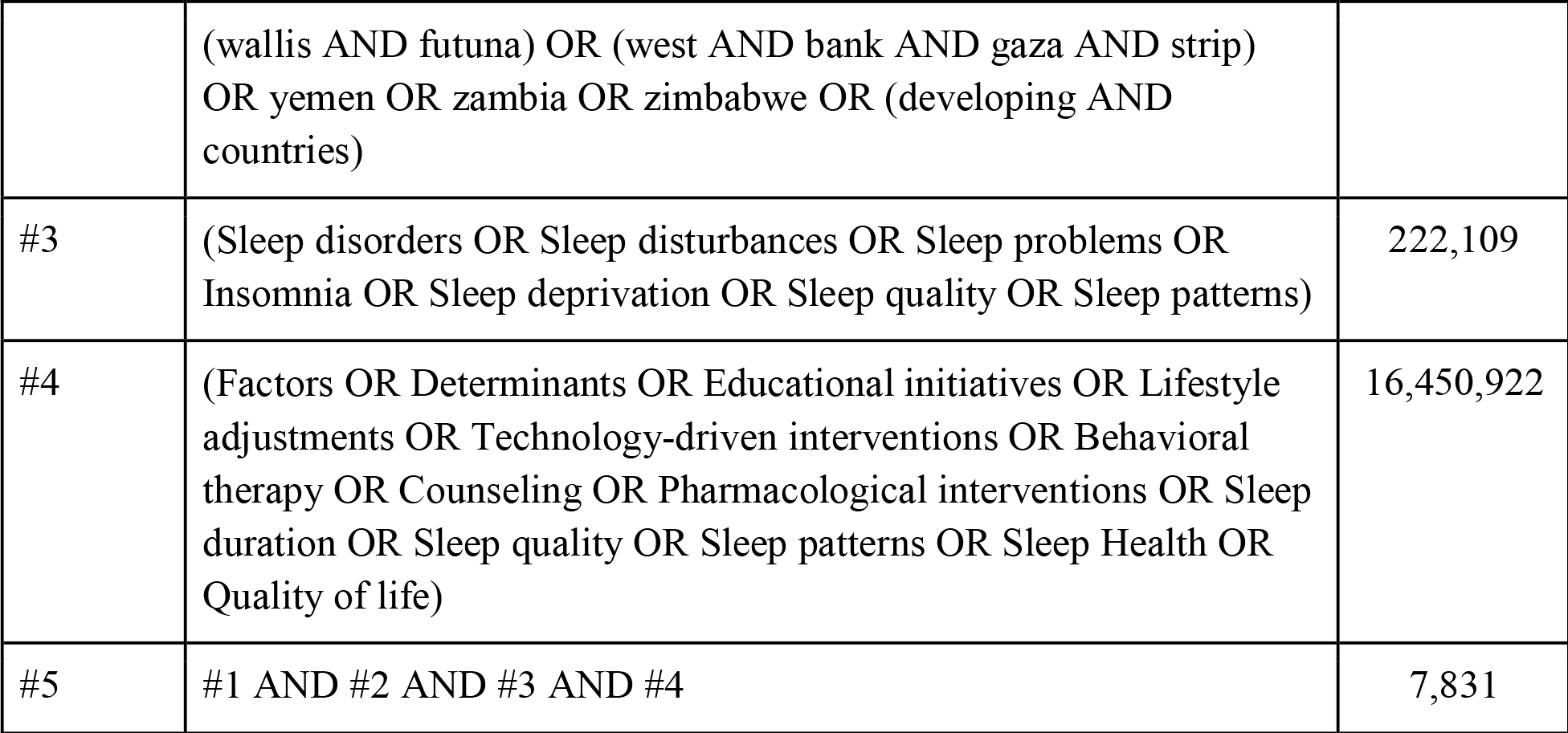

#### Scopus

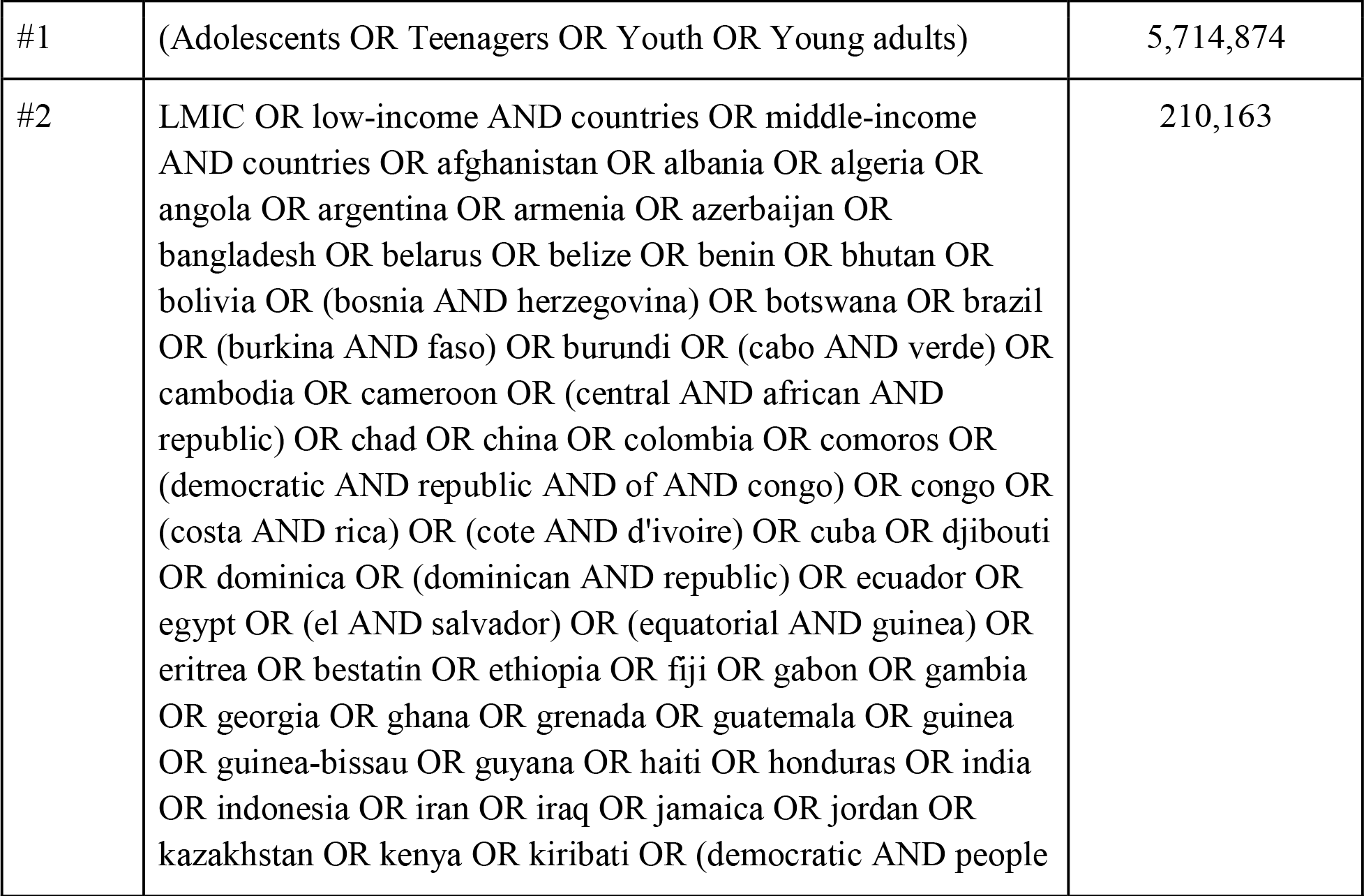
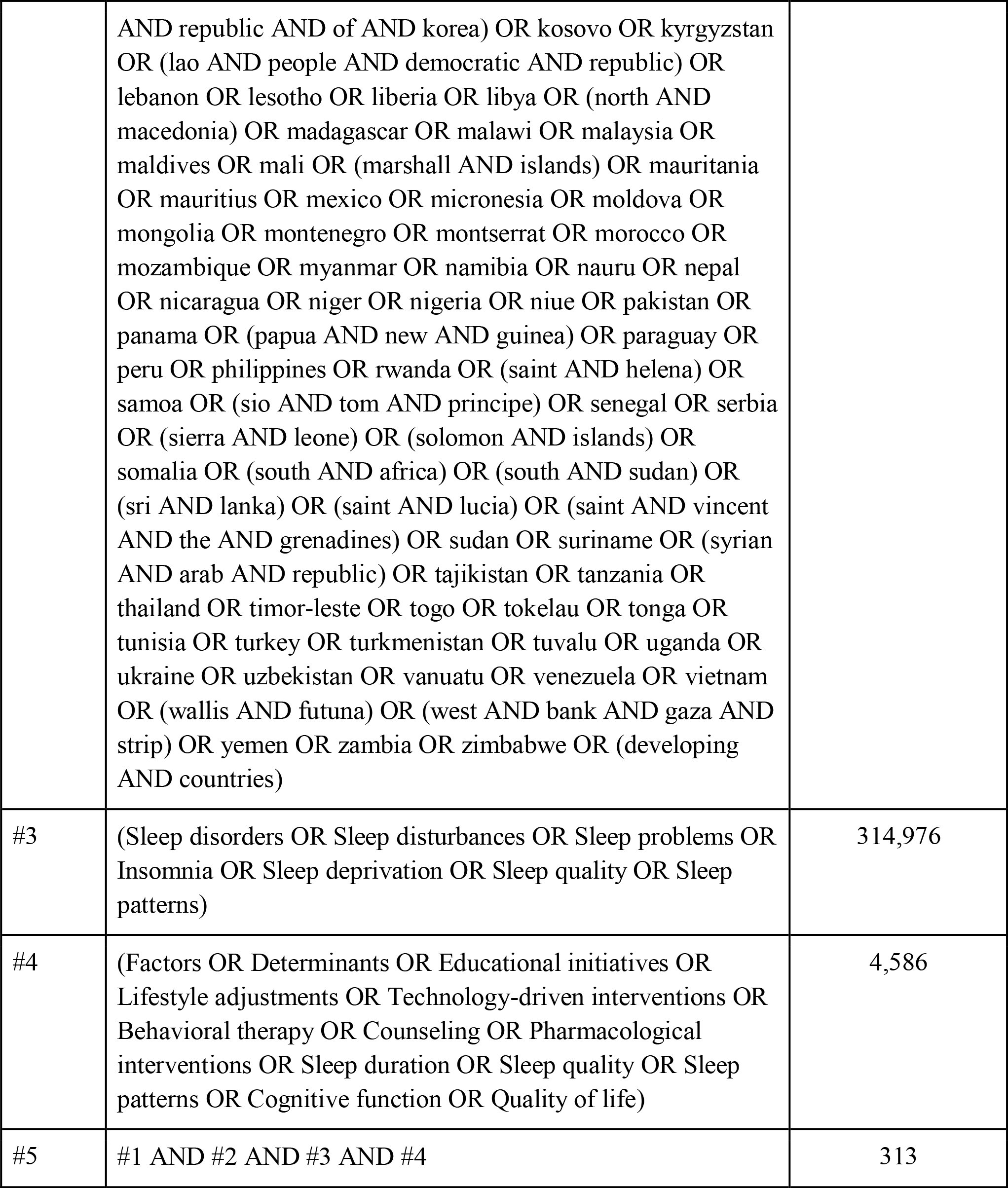

#### Embase

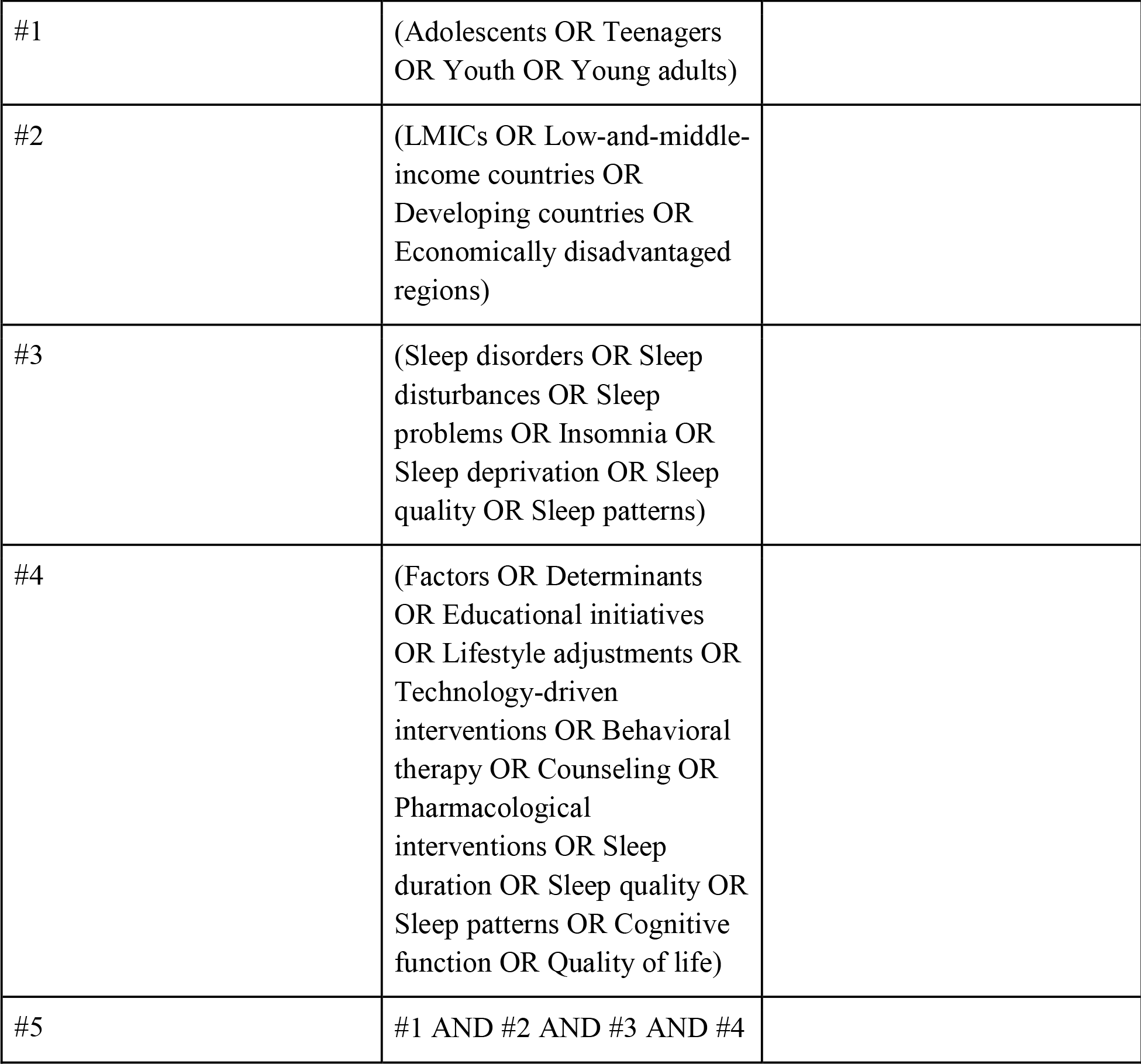

| Searches | Search strings | Results |
| --- | --- | --- |
| #1 | (Adolescents OR Teenagers<br>OR Youth OR Young adults) |  |
| #2 | (LMICs OR Low-and-middle-income countries OR<br>Developing countries OR<br>Economically disadvantaged<br>regions) |  |
| #3 | (Sleep disorders OR Sleep<br>disturbances OR Sleep<br>problems OR Insomnia OR<br>Sleep deprivation OR Sleep<br>quality OR Sleep patterns) |  |
| #4 | (Factors OR Determinants<br>OR Educational initiatives<br>OR Lifestyle adjustments OR<br>Technology-driven<br>interventions OR Behavioral<br>therapy OR Counseling OR<br>Pharmacological<br>interventions OR Sleep<br>duration OR Sleep quality OR<br>Sleep patterns OR Cognitive<br>function OR Quality of life) |  |
| #5 | #1 AND #2 AND #3 AND #4 |  |

#### PsychInfo

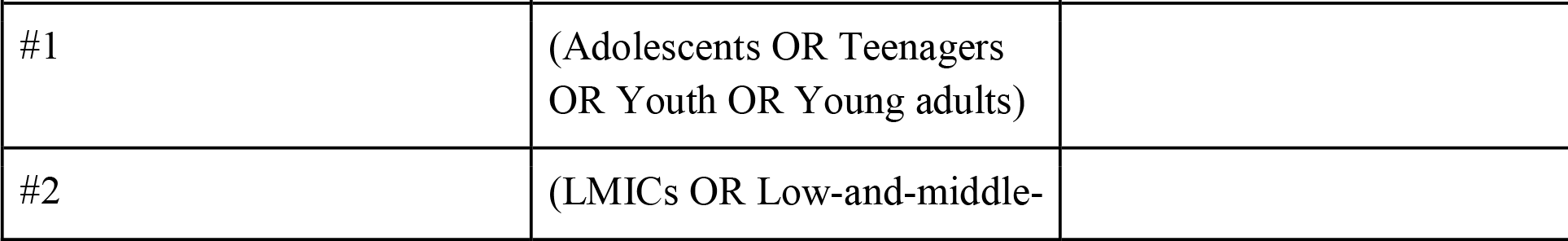
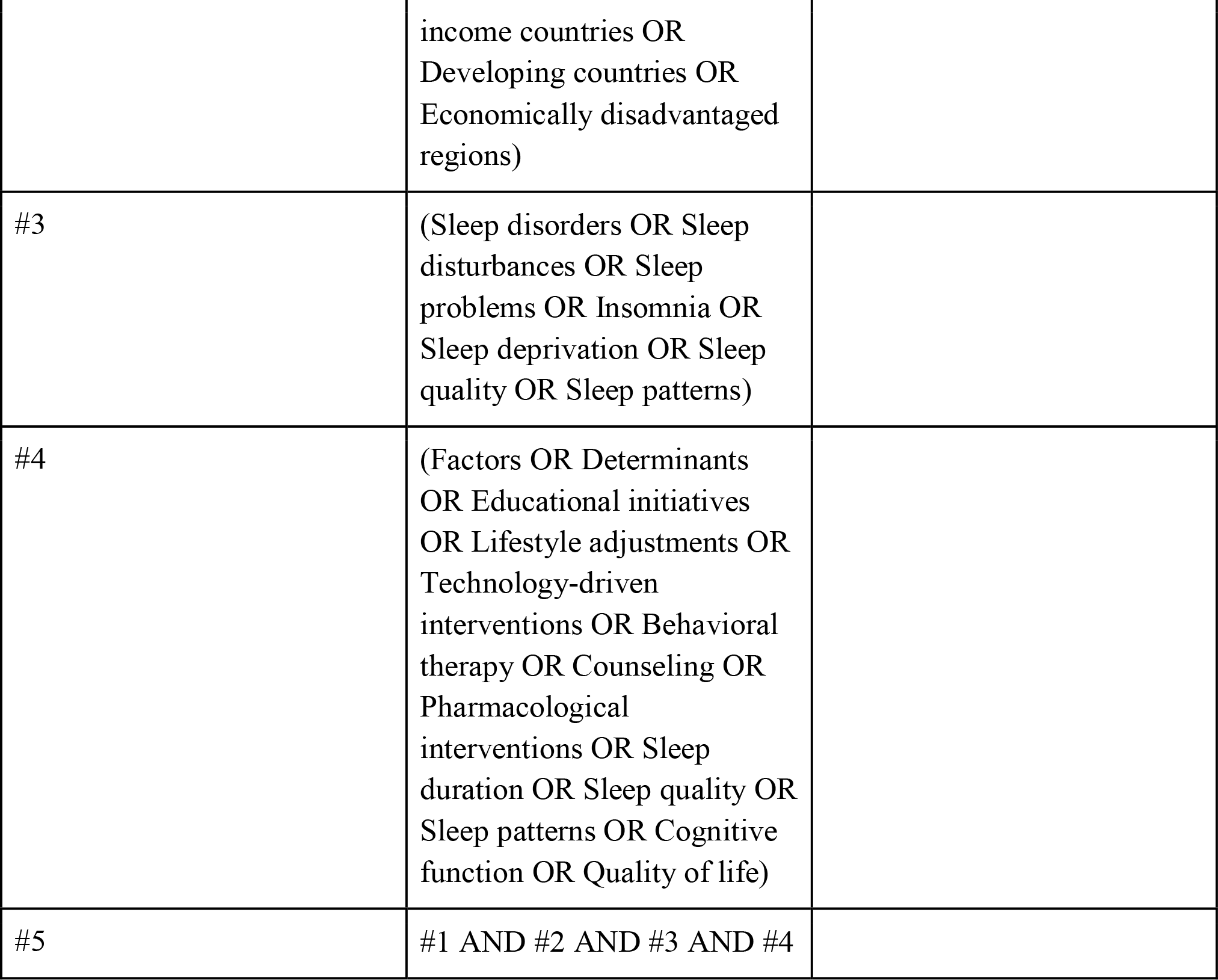

| Searches | Search strings | Results |
| --- | --- | --- |
| #1 | (Adolescents OR Teenagers<br>OR Youth OR Young adults) |  |
| #2 | (LMICs OR Low-and-middle- |  |
|  | income countries OR<br>Developing countries OR<br>Economically disadvantaged<br>regions) |  |
| #3 | (Sleep disorders OR Sleep<br>disturbances OR Sleep<br>problems OR Insomnia OR<br>Sleep deprivation OR Sleep<br>quality OR Sleep patterns) |  |
| #4 | (Factors OR Determinants<br>OR Educational initiatives<br>OR Lifestyle adjustments OR<br>Technology-driven<br>interventions OR Behavioral<br>therapy OR Counseling OR<br>Pharmacological<br>interventions OR Sleep<br>duration OR Sleep quality OR<br>Sleep patterns OR Cognitive<br>function OR Quality of life) |  |
| #5 | #1 AND #2 AND #3 AND #4 |  |

#### AJOL

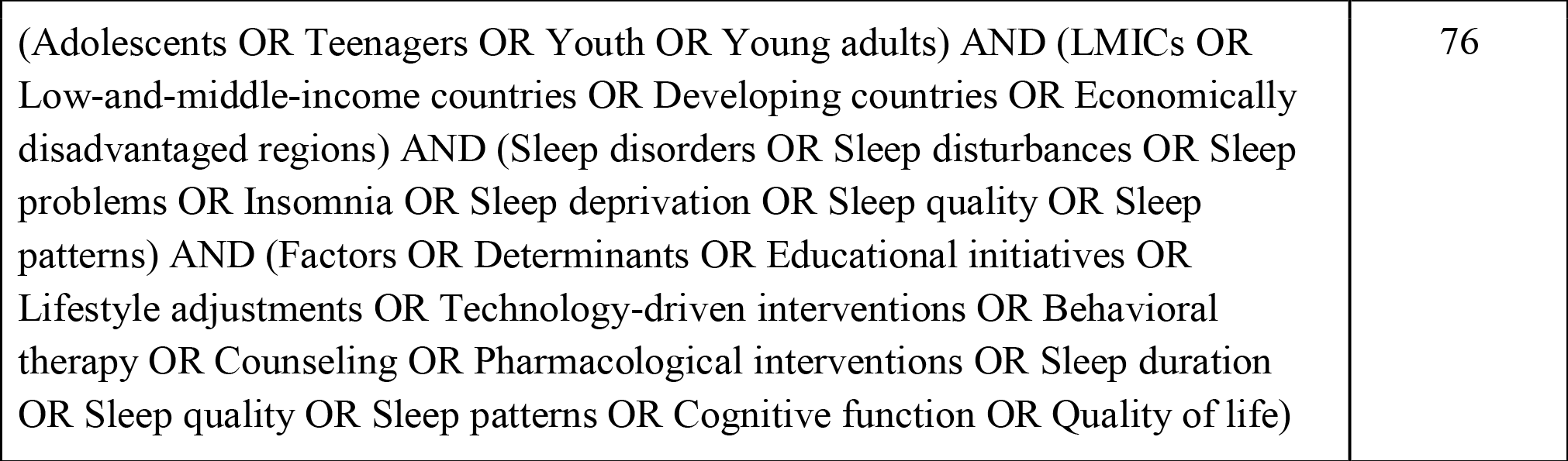

### Eligibility Criteria

#### Inclusion Criteria

- Original research studies, including randomized controlled trials (RCTs), clinical trials, and observational studies (cohort, case-control, and cross-sectional), assessing interventions to improve adolescent sleep health.
- Adolescents (aged 10-19 years) residing in low- and middle-income countries (LMICs).
- Studies evaluating various interventions include educational initiatives, lifestyle adjustments, technology-driven interventions, behavioral therapy or counseling, and pharmacological interventions.
- Studies that report quantitative outcomes related to sleep health (e.g., sleep duration, sleep quality, sleep patterns) in adolescents.
- Studies that were published from inception to date.
- Studies written in English or with available English translations from the journal

#### Exclusion Criteria

- Studies involving participants who are more than 19 years old.
- Animal studies, qualitative studies, review articles, case reports, and conference abstracts.
- Studies conducted only in high-income countries (HICs) or regions not classified as LMICs
- Studies published in languages other than English without available translations.
- Studies that focus on interventions unrelated to sleep health improvement in adolescents.

### Study Selection and Data Extraction

The researchers will employ a two-tier approach to ensure a meticulous selection of relevant studies. In the initial phase, two independent reviewers will comprehensively scrutinize the study titles and abstracts in the initial stage using our predefined eligibility criteria. In the subsequent stage, they will obtain and rigorously assess the full texts of studies identified as potentially relevant in the initial screening. In the event of conflicts about study eligibility, a third-party reviewer will resolve them.

The researchers will record and illustrate the transparency of our study selection process at different stages. The PRISMA-based flowchart will outline the excluded studies and comprehensively explain the reasons.

One author will extract the following information from all selected studies and organize the relevant data into a standard data extraction form. This form will include the following parameters for each study:

- General Information: Title, authors, publication date, study location, recruitment date, and study design
- Study Design: Information about sample size and follow-up.
- Participants: Inclusion and exclusion criteria, demographic particulars (age, gender, comorbidities, ethnicity).
- Intervention: Interventions used and the methods employed to assess those interventions.
- Outcomes: The level and various health-related outcomes, including sleep health, physical health, behavioral outcomes, psychological well-being, and other outcomes such as health-related quality of life.

### Data Synthesis

The researchers will synthesize data from the included studies through narrative synthesis and present findings in tables. If sufficient data are available and there is no significant heterogeneity, we will consider conducting a meta-analysis to compare outcomes across different categories.

### Assessment of Risk of Bias

For each outcome measure, two reviewers will comprehensively assess the biases independently. In the case of observational studies, we will employ the Newcastle-Ottawa Scale (NOS) to gauge study quality. Additionally, we will assess the randomized controlled trials (RCTs) using the Cochrane Collaboration risk of bias tool. Studies with a NOS score of ≥4 will be tagged as low risk of bias, rendering them eligible for inclusion in the systematic review. If there are any missing synthesis results, we will use funnel plots and Egger’s test to assess potential bias. We will use the Grading of Recommendations, Assessment, Development, and Evaluation (GRADE) approach to ensure certainty of the evidence for each outcome.

### Subgroup Analysis

If appropriate, we will perform subgroup analysis to explore the variations in the effectiveness of sleep health interventions across geographic regions with countries in LMICs and the impact on quality-of-life outcomes.

### Measurement of Effect

We will thoroughly analyze data obtained from qualified studies. This analysis will involve calculating odds ratios to assess the impact of various interventions on sleep health. We will calculate either the mean or standard mean difference for continuous, numerical, and categorical variables. Furthermore, we will also perform multivariate analyses to account for any potential factors that may introduce bias.

## Data Availability

All data that will be analyzed in this study is available at PubMed, Scopus, PsychInfo, and AJOL.

## Ethics and Dissemination

We intend to submit this study to a peer-reviewed academic journal and present our findings at conferences for broader dissemination.

## Authors’ Contributions

OEO & JOID wrote and revised the protocol draft for the final submission. TA wrote the Abstract while JOID edited it. All authors participated in the conceptualization of the research idea as well as proofreading the final manuscript.

## References

1. Brand S, Kirov R. Sleep and its importance in adolescence and in common adolescent somatic and psychiatric conditions. Int J Gen Med. 2011 Jun 7;4:425–42.

2. Gradisar M, Gardner G, Dohnt H. Recent worldwide sleep patterns and problems during adolescence: a review and meta-analysis of age, region, and sleep. Sleep Med. 2011 Feb;12(2):110–8.

3. Tarokh L, Saletin JM, Carskadon MA. Sleep in adolescence: physiology, cognition and mental health. Neurosci Biobehav Rev. 2016 Nov;70:182–8.

4. Neil-Sztramko SE, Caldwell H, Dobbins M. SchoolLbased physical activity programs for promoting physical activity and fitness in children and adolescents aged 6 to 18. Cochrane Database Syst Rev. 2021 Sep 23;2021(9):CD007651.

5. Wolfson AR, Harkins E, Johnson M, Marco C. Effects of the Young Adolescent Sleep Smart Program on sleep hygiene practices, sleep health efficacy, and behavioral well-being,. Sleep Health. 2015 Sep;1(3):197–204.

6. Inhulsen MMR, Busch V, van Stralen MM. Effect Evaluation of a SchoolLBased Intervention Promoting Sleep in Adolescents: A ClusterLRandomized Controlled Trial. J Sch Health. 2022 Jun;92(6):550–60.

7. Quante M, Khandpur N, Kontos EZ, Bakker JP, Owens JA, Redline S. A qualitative assessment of the acceptability of a smartphone app for improving sleep behaviors in low income and minority adolescents. Behav Sleep Med. 2019;17(5):573–85.

8. Adolescent-Sleep-Intervention Research: Current State and Future Directions - Matthew J. Blake, Melissa D. Latham, Laura M. Blake, Nicholas B. Allen, 2019 [Internet]. [cited 2023 Sep 25]. Available from: https://journals.sagepub.com/doi/10.1177/0963721419850169

