## Supplemental Tables for "Protocol for Systematic Review of Interventions to Improve Sleep Health in Adolescents Living in Low-and-Middle-Income Countries"

The researchers will identify eligible studies published from inception to date using an exhaustive search strategy across databases like PubMed, Google Scholar, Embase, Scopus, PsychInfo, and AJOL. This systematic review will follow the Preferred Reporting Items for Systematic Review and Meta-Analysis Protocols (PRISMA-P) guidelines.

The researchers will search the titles/abstracts/keywords using these search terms and boolean characters:

**PubMed**

| Searches | Search strings (24th September, 2023) | Results |
| --- | --- | --- |
| #1 | (Adolescents OR Teenagers OR Youth OR Young adults) | 3,026,051 |
| #2 | LMIC OR low-income AND countries OR middle-income AND countries OR afghanistan OR albania OR algeria OR angola OR argentina OR armenia OR azerbaijan OR bangladesh OR belarus OR belize OR benin OR bhutan OR bolivia OR (bosnia AND herzegovina) OR botswana OR brazil OR (burkina AND faso) OR burundi OR (cabo AND verde) OR cambodia OR cameroon OR (central AND african AND republic) OR chad OR china OR colombia OR comoros OR (democratic AND republic AND of AND congo) OR congo OR (costa AND rica) OR (cote AND d'ivoire) OR cuba OR djibouti OR dominica OR (dominican AND republic) OR ecuador OR egypt OR (el AND salvador) OR (equatorial AND guinea) OR eritrea OR bestatin OR ethiopia OR fiji OR gabon OR gambia OR georgia OR ghana OR grenada OR guatemala OR guinea OR guinea-bissau OR guyana OR haiti OR honduras OR india OR indonesia OR iran OR iraq OR jamaica OR jordan OR kazakhstan OR kenya OR kiribati OR (democratic AND people AND republic AND of AND korea) OR kosovo OR kyrgyzstan OR (lao AND people AND democratic AND republic) OR lebanon OR lesotho OR liberia OR libya OR (north AND macedonia) OR madagascar OR malawi OR malaysia OR maldives OR mali OR (marshall AND islands) OR mauritania OR mauritius OR mexico OR micronesia OR moldova OR mongolia OR montenegro OR montserrat OR morocco OR mozambique OR myanmar OR namibia OR nauru OR nepal OR nicaragua OR niger OR nigeria OR niue OR pakistan OR panama OR (papua AND new AND guinea) OR paraguay OR peru OR philippines OR rwanda OR (saint AND helena) OR samoa OR (sio AND tom AND principe) OR senegal OR serbia OR (sierra AND leone) OR (solomon AND islands) OR somalia OR (south AND africa) OR (south AND sudan) OR (sri AND lanka) OR (saint AND lucia) OR (saint AND vincent AND the AND grenadines) OR sudan OR suriname OR (syrian AND arab AND republic) OR tajikistan OR tanzania OR thailand OR timor-leste OR togo OR tokelau OR tonga OR tunisia OR turkey OR turkmenistan OR tuvalu OR uganda OR ukraine OR uzbekistan OR vanuatu OR venezuela OR vietnam OR (wallis AND futuna) OR (west AND bank AND gaza AND strip) OR yemen OR zambia OR zimbabwe OR (developing AND countries) | 6,809,569 |
| #3 | (Sleep disorders OR Sleep disturbances OR Sleep problems OR Insomnia OR Sleep deprivation OR Sleep quality OR Sleep patterns) | 222,109 |
| #4 | (Factors OR Determinants OR Educational initiatives OR Lifestyle adjustments OR Technology-driven interventions OR Behavioral therapy OR Counseling OR Pharmacological interventions OR Sleep duration OR Sleep quality OR Sleep patterns OR Sleep Health OR Quality of life) | 16,450,922 |
| #5 | #1 AND #2 AND #3 AND #4 | 7,831 |

**Scopus**

| Searches | Search strings (24th September, 2023) | Results |
| --- | --- | --- |
| #1 | (Adolescents OR Teenagers OR Youth OR Young adults) | 5,714,874 |
| #2 | LMIC OR low-income AND countries OR middle-income AND countries OR afghanistan OR albania OR algeria OR angola OR argentina OR armenia OR azerbaijan OR bangladesh OR belarus OR belize OR benin OR bhutan OR bolivia OR (bosnia AND herzegovina) OR botswana OR brazil OR (burkina AND faso) OR burundi OR (cabo AND verde) OR cambodia OR cameroon OR (central AND african AND republic) OR chad OR china OR colombia OR comoros OR (democratic AND republic AND of AND congo) OR congo OR (costa AND rica) OR (cote AND d'ivoire) OR cuba OR djibouti OR dominica OR (dominican AND republic) OR ecuador OR egypt OR (el AND salvador) OR (equatorial AND guinea) OR eritrea OR bestatin OR ethiopia OR fiji OR gabon OR gambia OR georgia OR ghana OR grenada OR guatemala OR guinea OR guinea-bissau OR guyana OR haiti OR honduras OR india OR indonesia OR iran OR iraq OR jamaica OR jordan OR kazakhstan OR kenya OR kiribati OR (democratic AND people AND republic AND of AND korea) OR kosovo OR kyrgyzstan OR (lao AND people AND democratic AND republic) OR lebanon OR lesotho OR liberia OR libya OR (north AND macedonia) OR madagascar OR malawi OR malaysia OR maldives OR mali OR (marshall AND islands) OR mauritania OR mauritius OR mexico OR micronesia OR moldova OR mongolia OR montenegro OR montserrat OR morocco OR mozambique OR myanmar OR namibia OR nauru OR nepal OR nicaragua OR niger OR nigeria OR niue OR pakistan OR panama OR (papua AND new AND guinea) OR paraguay OR peru OR philippines OR rwanda OR (saint AND helena) OR samoa OR (sio AND tom AND principe) OR senegal OR serbia OR (sierra AND leone) OR (solomon AND islands) OR somalia OR (south AND africa) OR (south AND sudan) OR (sri AND lanka) OR (saint AND lucia) OR (saint AND vincent AND the AND grenadines) OR sudan OR suriname OR (syrian AND arab AND republic) OR tajikistan OR tanzania OR thailand OR timor-leste OR togo OR tokelau OR tonga OR tunisia OR turkey OR turkmenistan OR tuvalu OR uganda OR ukraine OR uzbekistan OR vanuatu OR venezuela OR vietnam OR (wallis AND futuna) OR (west AND bank AND gaza AND strip) OR yemen OR zambia OR zimbabwe OR (developing AND countries) | 210,163 |
| #3 | (Sleep disorders OR Sleep disturbances OR Sleep problems OR Insomnia OR Sleep deprivation OR Sleep quality OR Sleep patterns) | 314,976 |
| #4 | (Factors OR Determinants OR Educational initiatives OR Lifestyle adjustments OR Technology-driven interventions OR Behavioral therapy OR Counseling OR Pharmacological interventions OR Sleep duration OR Sleep quality OR Sleep patterns OR Cognitive function OR Quality of life) | 4,586 |
| #5 | #1 AND #2 AND #3 AND #4 | 313 |

### **Prisma Flowchart**

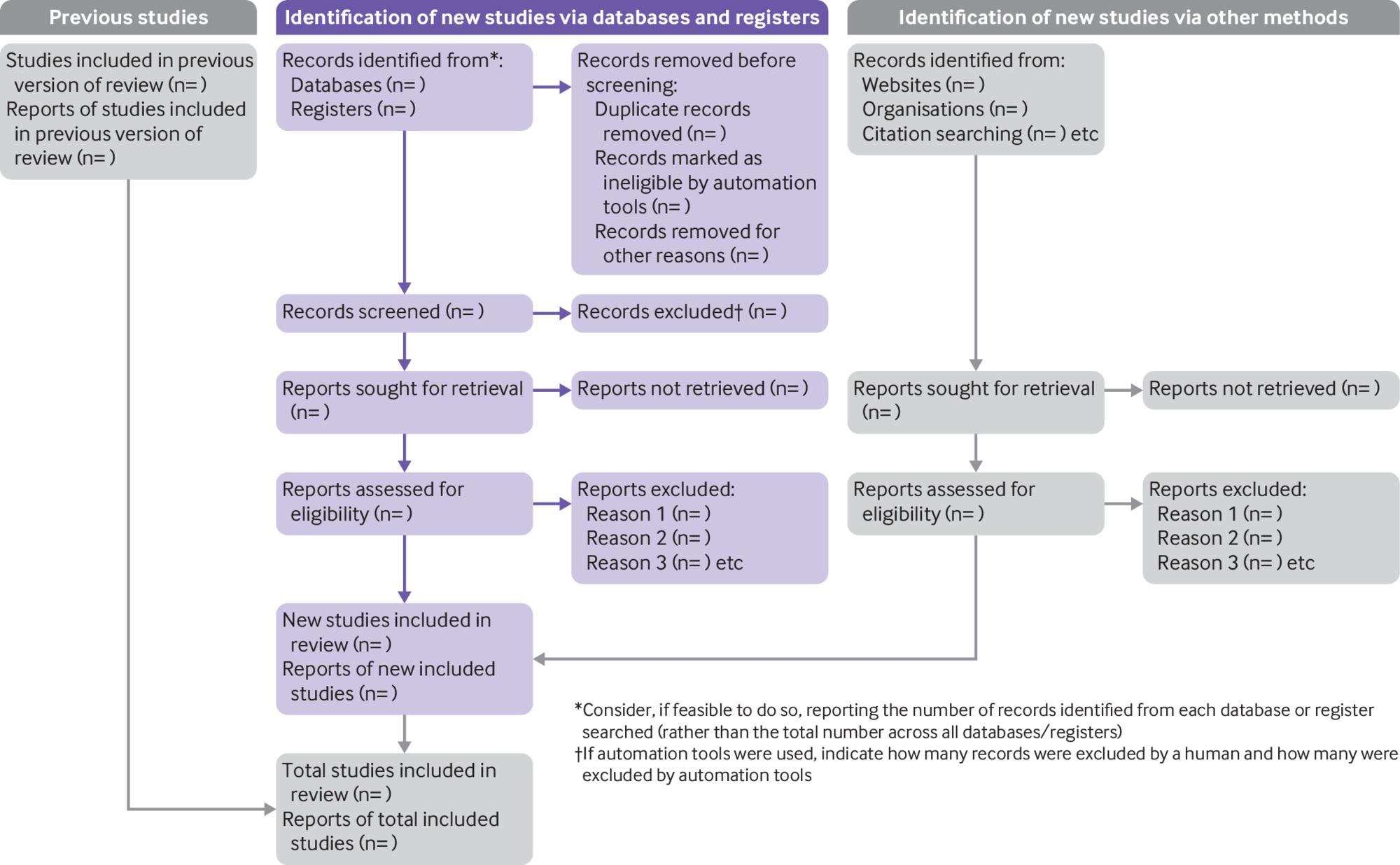

Flow chart: The PRISMA- P flow diagram shown will be used to illustrate the processes involved in conducting this systematic review.
